# AutoTriage - An Open Source Edge Computing Raspberry Pi-based Clinical Screening System

**DOI:** 10.1101/2020.04.09.20059840

**Authors:** Chaitra Hegde, Zifan Jiang, Pradyumna Byappanahalli Suresha, Jacob Zelko, Salman Seyedi, Monique A. Smith, David W. Wright, Rishikesan Kamaleswaran, Matt A. Reyna, Gari D. Clifford

## Abstract

With the recent COVID-19 pandemic, healthcare systems all over the world are struggling to manage the massive increase in emergency department (ED) visits. This has put an enormous demand on medical professionals. Increased wait times in the ED increases the risk of infection transmission. In this work we present an open-source, low cost, off-body system to assist in the automatic triage of patients in the ED based on widely available hardware. The system initially focuses on two symptoms of the infection fever and cyanosis. The use of visible and far-infrared cameras allows for rapid assessment at a 1m distance, thus reducing the load on medical staff and lowering the risk of spreading the infection within hospitals. Its utility can be extended to a general clinical setting in non-emergency times as well to reduce wait time, channel the time and effort of healthcare professionals to more critical tasks and also prioritize severe cases.

Our system consists of a Raspberry Pi 4, a Google Coral USB accelerator, a Raspberry Pi Camera v2 and a FLIR Lepton 3.5 Radiometry Long-Wave Infrared Camera with an associated IO module. Algorithms running in real-time detect the presence and body parts of individual(s) in view, and segments out the forehead and lip regions using PoseNet. The temperature of the forehead-eye area is estimated from the infrared camera image and cyanosis is assessed from the image of the lips in the visible spectrum. In our preliminary experiments, an accuracy of 97% was achieved for detecting fever and 77% for the detection of cyanosis, with a sensitivity of 91% and area under the receiver operating characteristic curve of 0.91. Heart rate and respiratory effort are also estimated from the visible camera.

Although preliminary results are promising, we note that the entire system needs to be optimized before use and assessed for efficacy. The use of low-cost instrumentation will not produce temperature readings and identification of cyanosis that is acceptable in many situations. For this reason, we are releasing the full code stack and system design to allow others to rapidly iterate and improve the system. This may be of particular benefit in low-resource settings, and low-to-middle income countries in particular, which are just beginning to be affected by COVID-19.

## Introduction

With a dramatic increase in the emergency department (ED) visit rates over the last four decades, both in the United States and around the world (1–4), accurate and timely triage is essential for assuring patients’ safety and optimal resource allocation. Crowding in the ED can affect the triage process, leading to longer waiting times for triage, longer ED length of stays, and potentially poorer outcomes (5). More acutely, crowding in ED during a pandemic such as COVID-19 could increase the risk for health professionals as well as patients.

Computer-aided triage systems have been proposed over the years with the help of browser-based applications that exchange information with existing medical records (6), wearable sensors (7) and automatic initial interpretation of CT scans (8). However, most existing methods either require significant interaction between the patients and the healthcare workers or need active input from the patients. Hence, there is an emerging need for an automatic triage system that works passively and requires minimal attention and interaction from both patients and health professionals. In this work, we focus on real-time identification of febrile status and cyanosis in patients, and estimation of heart-rate and respiratory effort.

The Emergency Severity Index (ESI), used by most EDs in the United States (9), records the febrile state of young children and the manifestation of cyanosis in all age groups. While core temperature is difficult to measure non-invasively, there is some evidence that infrared cameras can do so to some level of acceptable accuracy (10). In particular, we considered temperature in the forehead area and color distribution of the lip as indicators for the febrile state and cyanosis.

The Merck Manual (11) defines fever as an elevated body temperature that is higher than 37.8°C orally (12) and the Cleveland Clinic advised patients with a fever higher than 100.4°F / 38°C to isolate themselves as of April 2020 (13).

Cyanosis is a bluish discoloration of the skin or other areas of the peripheral body resulting from poor circulation or inadequate oxygenation of the blood. More specifically, it is due to an increased concentration of reduced hemoglobin (Hb) in the circulation and is clinically evident at an oxygen saturation of 85% or less. Mild cyanosis is more challenging to detect. Cyanosis can be observed in the lips, ears, trunk, nailbed, hands, and conjunctiva. Circumoral areas (around the mouth) have been compared in detecting cyanosis resulting from arterial hypoxemia. It has been noted that while the tongue is the most sensitive area, the lips are more specific ((14), chapter 45). For this work, we, therefore, focus on the lips, since they are easier to observe than the tongue and have been identified in ED emergency response assessment systems.

Assessing the heart rate is crucial in determining the overall health of an individual (15). Fever, which generally causes an elevated heart rate (16), fever is one of the key symptoms of coronavirus (17). While there is no standard for ‘high heart rate’ (that is no high enough to be tachycardia) for arousing suspicion of infection, somewhere between 100 and 100 beats per minute (bpm) is generally considered cause for concern. (Obviously this depends on recent activity, time of day, age, underlying health conditions, for example.)

Another symptom of coronavirus, as detailed by the Centers for Disease Control and Prevention (CDC), is shortness of breath or difficulty breathing (17). This results in abnormal breathing patterns. Estimating a metric that captures the respiration rate can help with classifying breathing as normal or abnormal, but is a rather brutal approach that can miss the difficulties encountered. A disordered respiratory effort may provide more useful information.

In this work we describe a system consisting of a low-cost minicomputer a Raspberry Pi, a Google Coral USB accelerator tensor processing unit (TPU), a visible light camera and a thermal camera, which are all portable and relatively inexpensive. By leveraging computer vision, signal processing, and machine learning classification techniques, the system is designed to be capable of segmenting out regions of interest and classifying the subject as febrile and/or cyanotic in real time (frame-by-frame). We also describe methods to estimate heart rate and respiratory effort in real-time using the same system.

## Related work

### A. Febrile state detection

Infrared imaging has been used extensively for remote, contactless human body temperature estimation for the last two decades since SARS (18). The efficacy of using infrared imaging for mass fever screening was first validated in (19), which demonstrated an ability to detect hyperthermia and a good correlation between thermal scanner readings and ear temperature. The efficacy of using a handheld FLIR350 camera (20) and the FLIRONE camera (which uses the Lepton sensor) (21) for febrile state detection were then validated. Recently, a deep learning face detection method was introduced to detect the face in thermal images for febrile state detection (22). However, the deep learning approach is only used intermittently due to the high processing time, and the region of interest was arbitrarily defined, leading to potential limitations in the usage.

### B. Cyanosis detection

Assessment of cyanosis is often conducted visually by doctors. The only known general method to semi-automatically detect cyanosis via a color correction and manual lips segmentation was proposed in a brief conference article (23).

### C. Heart rate estimation

Heart rate is one of the most frequently measured human vital signs. It is usually measured using electrocardiography, pulse oximetry, or by counting by radial palpation (24). With advances in the field of computer vision, various camera-based heart rate estimation methods have emerged. These methods have the advantage of being contactless. Subtle head motion is caused by the Newtonian reaction to the influx of blood at each beat. In (25), the authors report a method which leverages this behavior to measure the heart rate. The motion of the head is extracted by feature tracking and principal component analysis is applied to decompose the trajectories of the features into its component motions. The component which has a frequency spectrum that corresponds to the cardiac frequency interval is selected. The motion in this component is analyzed to identify peaks, which correspond to heartbeats. However, various noise sources such as internal and external head motions, low facial frame quality from video or camera and abnormal posture affect the heart rate estimation. Some solutions to these issues are proposed in (26), which introduces a face quality assessment method to ensure that low-quality frames do not contribute to the estimation of heart rate. Feature points from the face are combined with facial landmarks in order to create stable trajectories that are used to estimate heart rate.

The first remote photoplethysmogram (rPPG) imaging method, which used ambient light to estimate heart rate, was introduced in (27). The red, green and blue (R, G, and B) channels are extracted from a region of interest (ROI) in the frame. A raw photoplethysmogram (PPG) signal is generated using the spatial average pixel values of the channels from each frame over time. This raw signal is bandpass filtered to remove noise.

In (28) authors extract PPG signals from facial videos using blind source separation. The mean R, G, B channel values are calculated for the ROI in each frame over time. Here the ROI is the entire face. These raw signals generated from the means are normalized and decomposed into three source signals using Independent Component Analysis. The source signal which corresponds closest with a PPG signal is used to measure the heart rate, which somewhat limits the utility, since the heart rate estimation must be seeded with a known heart rate in the first place. A detailed review of many other heart rate estimation methods is given in (29).

### D. Respiratory effort estimation

From non-invasive methods to non-contact methods, various methods have been proposed to automatically estimate respiration rate and respiratory effort. Most early methods focused on replacing invasive methods like esophageal manometry via non-invasive methods, such as measuring the external breathing airflow via nasal cannula/pressure transducer system (30), measuring movement using diaphragm mechanomyography (31) or accelerometer (32), or measuring indirectly from surface electrocardiogram (33).

More recently, contactless approaches have gained popularity. A vast majority of them utilized either rPPG or a certain type of measurement for respiration induced motion. Karlen *et al*. acquired rPPG was via video in (34, 35) and proved to be feasible to estimate respiratory rates. However, respiratory effort estimation was not addressed in methods using the rPPG approach. The effectiveness of motion measurement based methods has also demonstrated with Doppler radar (36) and visible video (37). Additionally, it has been reported that respiratory pattern can be reconstructed with a motion based method (37), which enables estimation of respiratory effort.

Many other works have described attempts to derive physiology from video cameras, and an extensive comparison of the literature can be found in (38). In particular, the authors note that many approaches have been described in this field and that ‘the lack of standardization hinders comparability of these techniques and of their performance’. Notably they advise that sharing algorithms and/ or datasets would address this issue and potentially allow the application of newer techniques, such as deep learning’. Notably, none of the reviewed systems are available open source or have been tested in extremely large populations in noisy real-world settings. In this work we leverage deep learning algorithms developed on public data, and begin the work of applying these using a framework that we have publicly released to help reach critical mass with public data and repeatable techniques.

## Methods

### E. Hardware configuration

The system proposed is an off-body camera based system to detect symptoms of respiratory illnesses. A Raspberry Pi 4 (RasPi) with 4GB RAM is used as the main processor and is used to run most of the algorithms. A Google Coral USB Accelerator is used to perform person detection, which uses a deep learning based algorithm, and thus needs significantly more computational power. The accelerator is designed to run deep learning models optimally.

A Raspberry Pi camera v2, which is a visible light camera, is used to detect people and for cyanosis, heart rate, and respiration effort estimation. A FLIR Lepton 3.5 Radiometry Long-Wave Infrared Camera with its associated IO module is used for febrile state detection. The left panel in Figure 1 shows the proposed system. An optional temperature and humidity sensor is added to help with the thermal camera calibration. For this work, we tested the Gowoops 2 PCS DHT22 Temperature Humidity Sensor Module. An optional car battery power-source is also detailed in this work for use in remote and rugged locations.

**Fig. 1.**
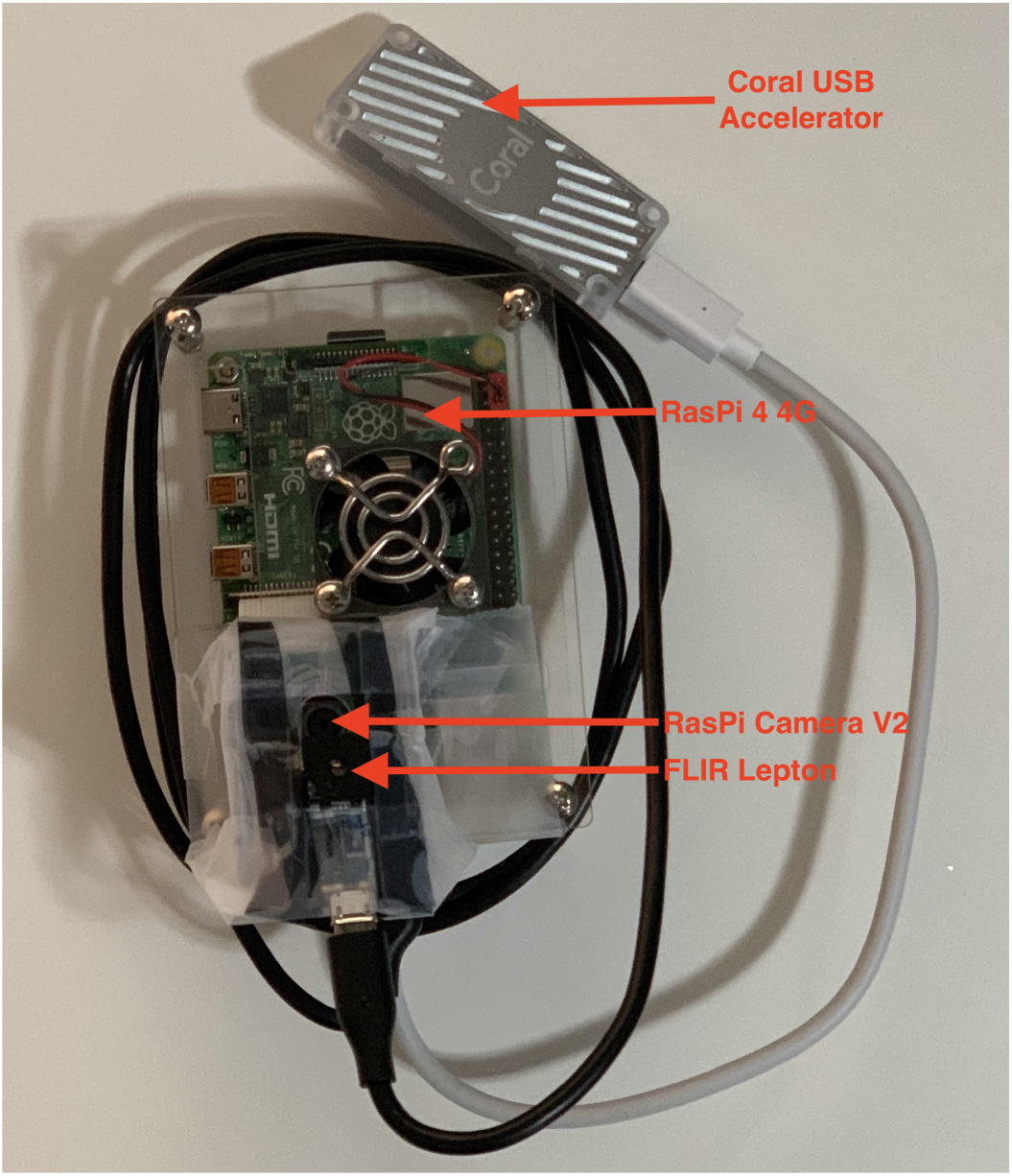
Left: Hardware configuration consisting of a Raspberry Pi 4 (4 GB RAM), a Google Coral USB accelerator (top), a RasPi Camera v2 and a FLIR Lepton 3.5 Radiometry Long-Wave Infrared Camera. Right: Installation of system with all cabling including power and external monitor for visualization (not shown or needed for detection).

Our system works in real-time, with the visible light camera capturing frames at a rate of 25Hz and the thermal camera at a rate of 9Hz. The various estimation results can be displayed on an external monitor via an HDMI-microHDMI cable or on an LCD display screen directly connected to the RasPi. (We chose a $20 touch-screen 3.5 inch LCD for compactness.) The full setup can be seen in the right panel of Figure 1. The costs associated with various parts are included in the appendix. The total retail off-the-shelf cost of the system is around $480 (not including the car battery).

### F. Electrical supply

The design of this system focuses on low-resource mass-triage scenarios, where power is limited or absent. We therefore chose to power our system from a car battery, which is a readily available power source across the planet. Since car batteries supply energy at 12V, a stepdown transformer is required. We chose a 12V to 5V DC step-down power converter that comes with a 15W output (3A at 5V). The transformer was equipped with a USB Type-C power supply output, which was connected to the RasPi.

Noting that the average car battery supplies 40 Ampere-hours or 144, 000 Coulombs at 12V for one charge cycle, and the RasPi together with the coral, transformer and LCD touchscreen draw 3A at 5V, the car battery can supply power for 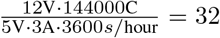 hours continuously before discharging. Assuming we do not want the car battery to go below 50% of its capacity, we expect the car battery to run the unit for 16 hours continuously before needing to be recharged.

### G. Temperature and Humidity Detection

A temperature-humidity detection module was added to assist with the calibration of the FLIR lepton camera. Specifically, we used the DHT22 sensor module that is comprised of a capacitive humidity sensor and a thermistor that measures the surrounding air to provide calibrated temperature and humidity values.

This module comes with a digital board that houses three pins, namely VCC, GND, and OUT. The sensor has an operating voltage of 3.3*/*5V (DC), and the OUT can be read from a GPIO pin on the RasPi. The temperature range is −40 to 80°C, and the humidity range is 0 *−* 100%RH. The associated Python software that allowed us to integrate the sensor with the RasPi is open source and available in our Github repository (39).

### H. Face and thorax detection

To estimate febrile state, cyanosis, heart rate, and respiration effort, it is necessary to detect people in a frame and segment out certain regions of the face. For febrile detection, the forehead-eye region is necessary. Heart rate estimation is performed on the area of the face below the eyes. For cyanosis estimation, the lips need to be segmented, and the thorax region is necessary for respiratory effort estimation.

We use PoseNet (40) to detect people in a frame. This is a convolutional neural network based algorithm which regresses *keypoints* of human beings in an image or video. Here, keypoints refer to image coordinates of certain key parts of the body, such as the elbows, knees, eyes, nose, etc.

We use the estimated keypoints of the left and right eyes to extract the various face segments mentioned above. If we define the distance between the eyes to be D pixels, then to determine the bounding box around the forehead-eye region, we use a rectangle that has width 2D and a height 1.2D. The base of the bounding box is 0.2D below the eyes. This ensures that the forehead-eyes area is captured. This is an important site since the inner canthus of the eye is consistently the warmest area on the head and the most suitable area for fever detection (41). To create a bounding box around the lips, we move a distance D below the eyes and create a bounding box with width D and height 0.5D. For heart rate estimation, the bounding box has a height of 0.8D, starting 0.2D below the eyes and extending downwards. It has width 1.2D, which extends from 0.1D left of the left eye to 0.1D right of the right eye. The bounding box for respiratory effort estimation is obtained by using the shoulder keypoints as reference. The top edge of the bounding box is formed by joining the shoulder keypoints. If we let the number of pixels between the shoulders be denoted, R, then the bottom edge is 1.5R pixels below the top edge. I.e., the bounding box has a width of R pixels and height of 1.5R pixels. Note that the coordinates obtained by applying this heuristic are rounded to the closest integer value

These values were set empirically (through trial and error using the authors as test points). With few subjects in a given lockdown space, large scale experiments in a short period of time were not possible. In future experiments, these values can be optimized on larger datasets. Figure 2 shows an example of the forehead and lip detection on one of the authors using this approach in the visible spectrum (upper plot) and the corresponding FLIR image (lower plot). Note that the images have slight FOV, image angle, and translational differences since the cameras cannot take images from the same location in space and operate at different (non-synchronous) sampling rates.

**Fig. 2.**
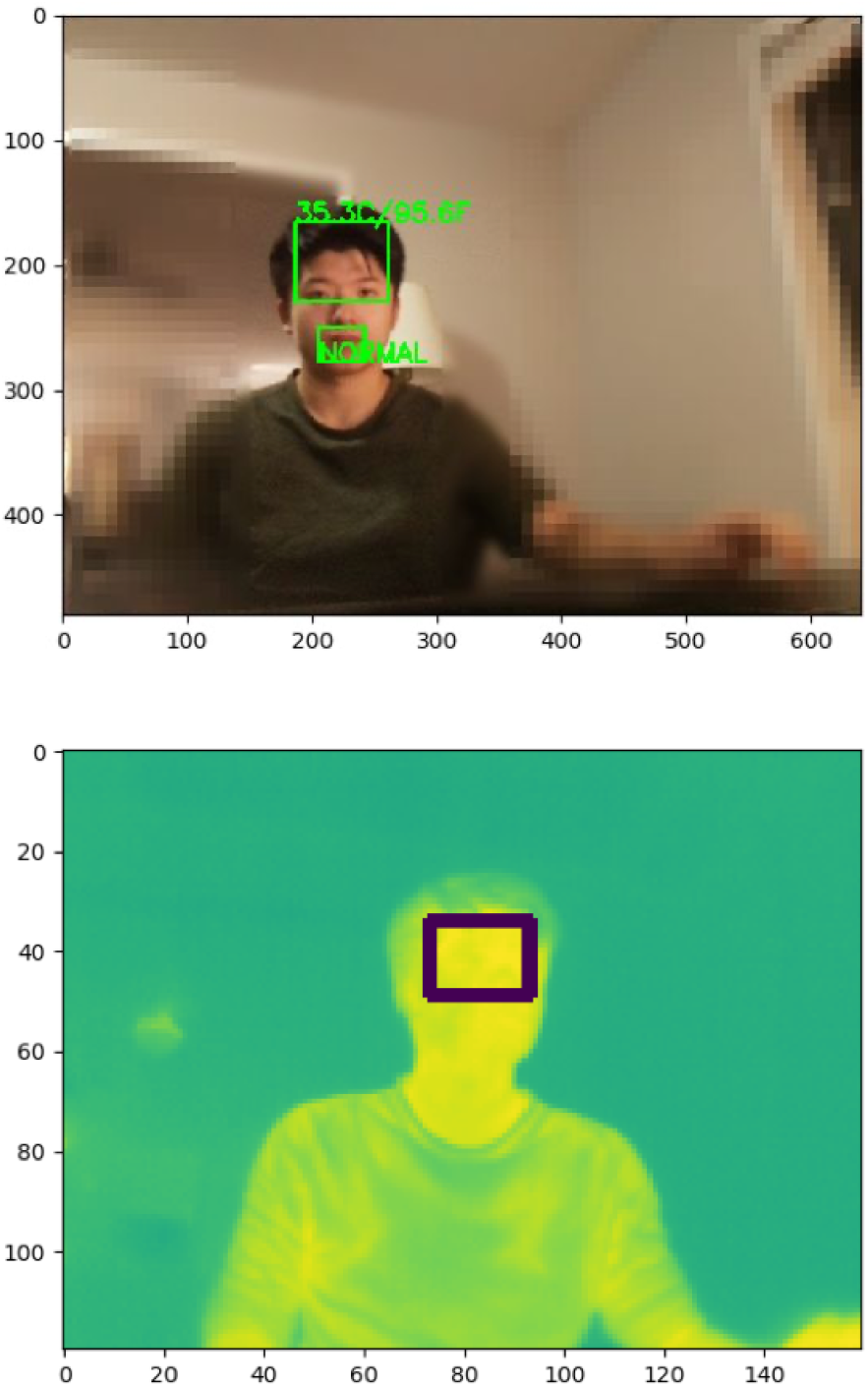
Bounding boxes detected for the forehead and lips from a 1m range in visible light video (upper image) and thermal video (lower image) using PoseNet.

### I. Febrile state detection

Unlike the previous studies, our proposed system detects key points using visible light video and then transforms the coordinates of the bounding boxes of the ROI to coordinates in the thermal video. After finding the ROI in the thermal video, the ten pixels exhibiting the highest temperatures are averaged to produce a final temperature estimate. Lastly, a threshold is set to determine the febrile state.

#### I.1 Thermal output calibration

To achieve a more accurate measurement of the body temperature, we followed the guidelines from the FLIR Lepton 3.5 datasheet (42) and used a robust regression to map the temperature output to the ground truth within the desired range.

We used bottles (with open lids) of heated water with temperature ranging from 35− 40°C as a heat source and located them at one meter to the camera and approximately in the center of the field of view (FOV). (See figure 3.)

**Fig. 3.**
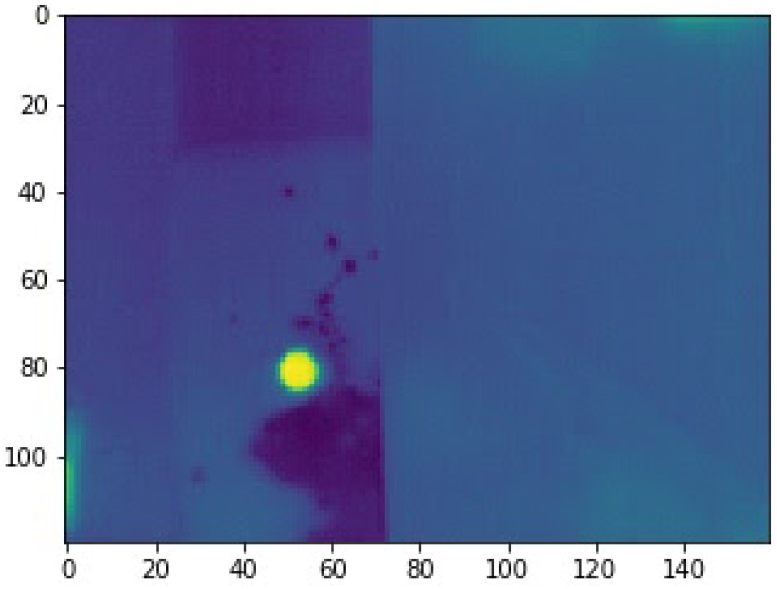
Illustration of the water calibration experiment. The bright spot in the lower left quadrant of the FLIR output represents the water heated to varying known temperatures.

The reference temperature of the water was measured three times using a Braun IRT6500 thermometer and averaged. The reference has an accuracy of ± 0.2°C within the 35 − 42°C measurement range. Also, the top ten pixels in the heat source were selected and averaged as the final output of the FLIR Lepton camera. The slope and the intercept are then fitted using the above-described experiment via Huber regression implemented in *scipy* 1.2.3. The Root Mean Square Error (RMSE) metric was used to evaluate the fitting error. The fitted line was then implemented to convert pixels values to temperature output.

#### I.2 ROI registration in thermal video

Since the forehead-eyes area is detected in the visible light image sequences, a coordinates transformation is needed to find the forehead-eyes location in the thermal video. The transformation depends on the resolutions and FOVs of the two cameras and the relative physical displacement between them. The resolution of the RasPi Camera is set at 1640 × 1232 pixels and the corresponding FOV is 62.2°horizontally and 48.8°vertically. The resolution of the FLIR Lepton camera is set to be 160 × 120 pixels and the corresponding FOV is 57°horizontally and 71°diagonally. Since there only exist less than 2 cm distance and consequently a small angle difference between the cameras, the transformation can be expressed as:

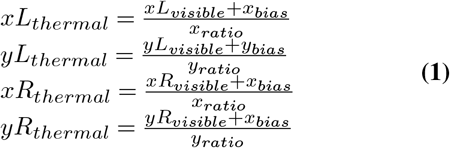

Where *xL, yL* are the coordinates for the left vertex of the bounding box and *xR, yR* denote the right vertex. *x*_*ratio*_, *y*_*ratio*_ are the resolution ratios between RasPi Camera and FLIR Lepton Camera. And *x*_*bias*_, *y*_*bias*_ are the view difference caused by different FOVs and can be calculated as:

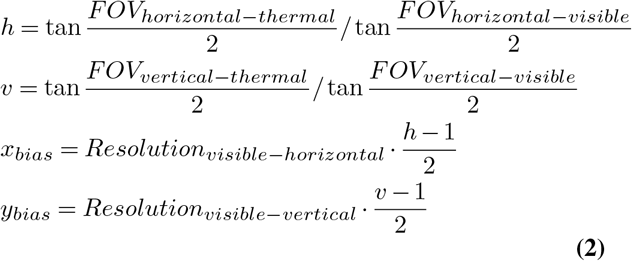

In practice, because of the angle difference and distance between the cameras, which varies with different mounting schemes, empirical offsets were added to ensure accurate transformation.

#### I.3. Threshold selection

Forehead (temporal) temperature is usually 0.5°F (0.3°C) to 1°F (0.6°C) lower than an oral temperature measurement (43). Combining the guideline from Cleveland Clinic (13), a threshold of 37.4°C was selected.

### J. Cyanosis detection

To detect cyanosis, we use the image of the lips, which is segmented as explained in section H. In this work, we have tested three classification algorithms: K nearest neighbor algorithm, logistic regression, and support vector classifier on a dataset of cyanotic and non-cyanotic lips. We picked the algorithm with the best performance to run in real-time on the RasPi. This algorithm classifies images of lips segmented from the frames coming from the visible light camera as cyanotic or non-cyanotic.

#### J.1. Dataset of cyanotic lips

A small dataset was created using images available on the internet. Images of cyanotic lips were identified using Google Image search, and those images with the word “cyanosis” in the description were chosen to be included in the dataset. Similarly, images of non-cyanotic or healthy lips were chosen from Google Images at random. (Human over-read was used to ensure quality.) These images were cropped to include only the lips and exclude other parts of the face. The dataset is balanced with 35 cyanotic lip images and 35 non-cyanotic lip images. We have attempted to make the dataset as race and age inclusive as possible. The non-cyanotic lip images are a mix of different races and also include a range of ages, from infants to elderly people. Finding such a mix of races was challenging for cyanotic lip images using internet-based image searches, which is consistent with the racial bias observed in other datasets (44). Approximately 91% of the cyanotic images belong to fair skinned people. The dataset is included in our Github repository (39). It is important to note that the labels for these images are not verified by independent healthcare experts. We assume that the description of the images that were available on the internet are correct.

#### J.2. Classification algorithm

The task of classifying lip images as cyanotic or non-cyanotic is a binary classification problem. As stated above, we implemented three binary classifiers for this purpose: K nearest neighbors (KNN), logistic regression (LR), and support vector classifier (SVC). We computed the frequency of pixel intensities from each color channel (R, G, B channels) and used this as input to the classifiers. A simple histogram with eighteen equally spaced bins was used for this purpose. The number of bins (i.e., 18) was a hyperparameter that was tuned. The rationale behind this was that the color distribution would be different in cyanotic and non-cyanotic lips, but there would be some colors in common. In other words, not all of the lip would be cyanotic, and some areas outside of the lip would be included. The bins representing these colors could then be regularized out.

For the KNN algorithm, we used *K* = 3 neighbors (chosen by hyperparameter tuning) and the Euclidean distance metric to calculate the distance between the neighbors. Logistic regression is a well known binary classification algorithm. In our implementation, we combined this with *L*_2_ regularization. For the SVC, a regularization parameter *C* = 2 was chosen via grid search. Radial basis function kernel was used to make SVC a non-linear classifier. Leave-one-out cross validation method was used for all three classifiers to report the results.

For all the three implementations above, we have used the *scikit-learn* package (version 0.22.2) (45) in Python 3.7.3. We have used the default *scikit-learn* values for the parameters which are not mentioned above.

### K. Heart rate estimation

Our implementation of heart rate estimation uses the visible light camera in our system. We have followed the algorithm detailed in (46) for our implementation. This method attempts to nullify the effects of the motion of the subject and illumination variation while measuring heart rate, which are common problems in a real-world setting. The steps involved are detailed below. It is worth noting that a similar pipeline can be applied to thermal video as well, and the fluctuation of the output in ROI can also be viewed as remote PPG.

#### K.1. ROI detection and tracking

In their original work, (46) use the Viola-Jones face detection algorithm (47) to detect faces in the frames. They then use the Discriminative Response Map Fitting method (48) to find facial landmarks to select the ROI, which is the area of the face below the eyes. This reduces interference caused by blinking and other eye movement and also excludes non-face pixels like the background.

In our implementation, we use PoseNet (40) to detect faces and identify the region of the face below the eyes, as explained in H. The Viola-Jones algorithm relies on the presence of eyes and the face looking straight into the camera in order to detect the face. If the face is at an angle to the camera, detection is not guaranteed. PoseNet does not suffer from this problem. The process of obtaining the ROI using PoseNet is explained in section H.

#### K.2. Illumination rectification

This step is performed in order to reduce the effect of illumination changes that occur in the environment where the system is placed. To achieve this, the authors in (46) calculate the mean value of all pixels in the green channel for the ROI extracted above and for the background region. Some amount of the mean green channel value of the background is subtracted from the face region. This amount is determined iteratively using a Normalized Least Mean Square adaptive filter (49). The authors use the Distance Regularized Level Set Evolution method (50) to segment out the background in order to obtain the mean green channel value. We use a simpler method that involves extracting pixels that are not part of the ROI and finding their mean green channel value. Note that the green channel is used over other channels since it contains the strongest plethysmographic signal out of all the three channels (27).

#### K.3. Non-rigid motion estimation

Non-rigid motions inside the ROI, such as small changes in expression, can disturb the heart rate estimation. To address this, the authors of (46) segment the illumination rectified mean values into smaller pieces and estimated their standard deviations. 5% of the segments with the highest standard deviations are excluded from further processing. In our implementation, we find that excluding 20% of segments with the highest standard deviation provided more accurate results during initial optimization.

#### K.4. Temporal filterin

Three filters are used by (46) on the signal obtained from the above step to exclude frequencies outside the range of interest. First, a detrending filter is used to remove the slow and non-stationary trend in the signal. Next, a moving average filter is used to remove random noise. Finally, a Hamming window-based finite impulse response bandpass filter is used to limit the signal to have frequencies in the desired range. The filter has cutoff frequencies of 0.7Hz and 4Hz to cover the normal range of heart rate from 42 beat-per-minute (bpm) to 240 bpm. After filtering, the power spectral density (PSD) of the signal is found using Welch’s method (51). The frequency with the maximum power is multiplied by 60 to obtain the heart rate in beatsper-minute.

### L. Respiration effort estimation

Respiration rate is a difficult parameter to estimate, given that it can be non-periodic, spectral estimation techniques are likely to fail. In particular, we are looking to observe the pattern of respiration (i.e., struggling to breath), rather than any respiratory rate (although hyperventilation is important to identify). Rather than estimating a rate, we chose to derive the respiratory effort tracing that can then be used to derive further metrics (e.g., high sample entropy with a peak in energy between 0.05-2 Hz may identify disordered, but rapid, breathing). Of course, much more data is needed before reliable metrics and thresholds can be determined.

To determine the respiratory effort, we isolate the upper thorax using a bounding box that covers the regions from the shoulders to the diaphragm. Then we calculated the first difference between each pixel in subsequent frames. The final effort signal is then given as the average of each ‘difference’ image after a post-processing bandpass filter step (with a passband of 0.5Hz and 2Hz).

## Experiments

### M. Febrile state detection

The preliminary test was conducted on a healthy male subject. A heated cloth was put on the subject’s forehead to raise the temperature of the subject’s forehead. The estimated temperature from the proposed system was recorded immediately after a measurement from the thermometer in the center of the subject’s forehead. The subject was sitting one meter away from the cameras. The RMSE between the estimated value and temperature from the thermometer was used to evaluate the accuracy of our proposed system.

### N. Heart rate estimation

To test the algorithm in real time, one of the authors was used as the subject. The subject was seated in an indoor setting at distances of 50cm and 1m from the system. The heart rate was measured using the system, and the pulse reading was taken at the wrist simultaneously. The pulse readings are used as ground truth. Each reading was one minute long. This was repeated for different heart rate by increasing the heart rate using cardiovascular exercises. The subject tried to stay as still as possible to get accurate readings. 30 readings were recorded in total, with 15 readings recorded at a distance of 50cm from the system and 15 readings recorded at a distance of 1m from the system.

### O. Respiratory effort estimation

To test this concept, one of the authors was used as a subject. The subject stood 0.5m away from the system, and the algorithm was run to estimate respiratory effort for 30 seconds in each trial. Respiratory rate was recorded simultaneously by counting the number of breaths per minute. There were a total of 14 trials where 7 were normal breathing, and the other 7 were faster and harder breaths to simulate shortness of breath. It was ensured that there was minimal illumination variation in the environment in order to avoid interference from varying ambient lighting.

## Results

### P. Febrile state detection

The left subfigure in figure 4 shows the measurements used in the calibration process and the fitted results. The calibrated values of the slope and intercept were determined to be 0.0113 and 313.383, respectively, and the resultant RMSE was 0.57°C. The right subfigure in figure 4 shows the measured data points in the preliminary experiment, in which the proposed system achieved an RMSE of 0.41°C and a Pearson correlation coefficient of 0.96. When applying 37.4°C as the threshold for febrile state detection, an accuracy of 96.7% and an area under receiver operating characteristic curve (AUC) of 0.97 were achieved.

**Fig. 4.**
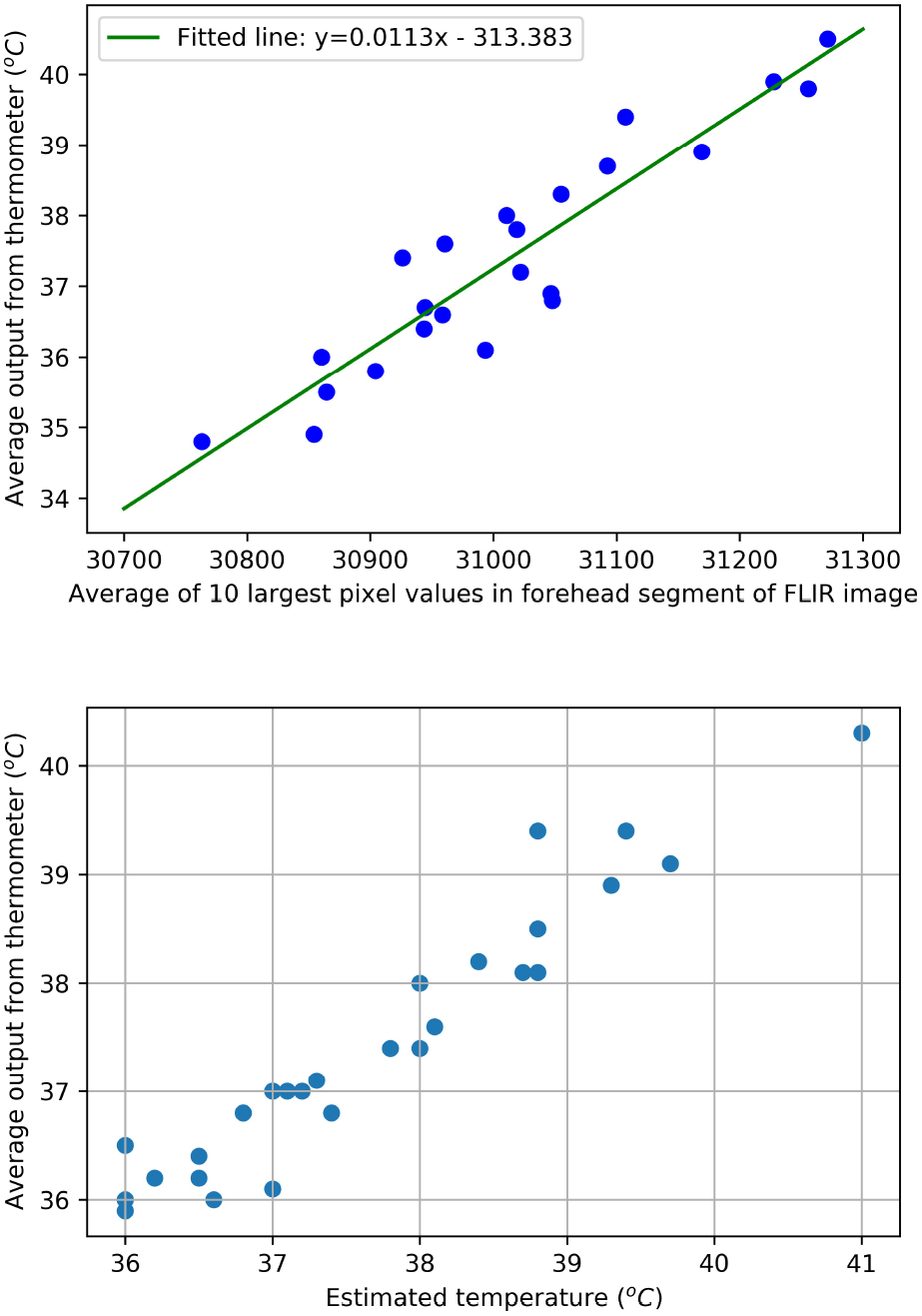
Upper: Calibration of FLIR images using a bottle of water: Best fitted line between the average temperature from the thermometer and the FLIR pixel values had a root mean square error (RMSE) of 0.57 °C. Lower: Estimated temperature vs. temperature measured from thermometer. The Pearson correlation between the parameters was found to be 0.96 and the RMSE difference between them was 0.41 °C.

Since the system described here was only created in an apartment with a narrow temperature and light range, which is not necessarily reflective of how this tool might be used in a real triage situation, we evaluated the system in Emory’s ED and discussed its utility with the ED team. When used in a bright cold environment, the temperature estimation appeared to run about 2 degrees °C lower than in the training environment, and the lip analysis triggered many false positives.

### Q. Cyanosis detection

Tables 1 shows the confusion matrices for each classifier. Figure 5 shows the receiver operating characteristic curve and AUC of each classifier. Table 2 summarizes the accuracy, AUC, sensitivity and specificity of each of the three classification models.

**Table 1.**
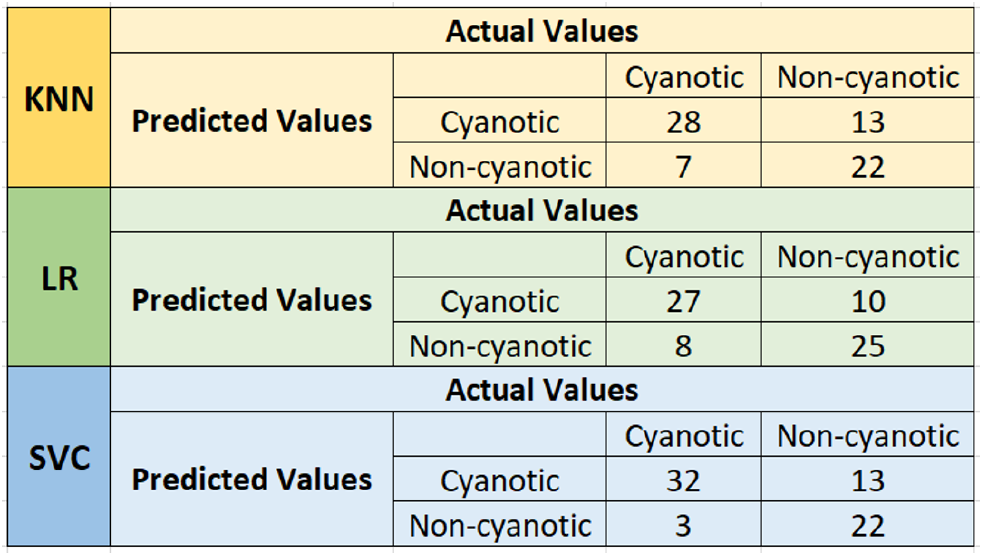
Confusion matrix for KNN, LR, SVC.

**Table 2.** Performance of assessed cyanosis detection classifiers.

| Algorithm | Accuracy (%) | AUC | Se | Sp |
| --- | --- | --- | --- | --- |
| KNN | 71.4 | 0.76 | 0.83 | 0.63 |
| LR | 74.3 | 0.73 | 0.77 | 0.71 |
| SVC | 77.2 | 0.83 | 0.91 | 0.63 |

**Fig. 5.**
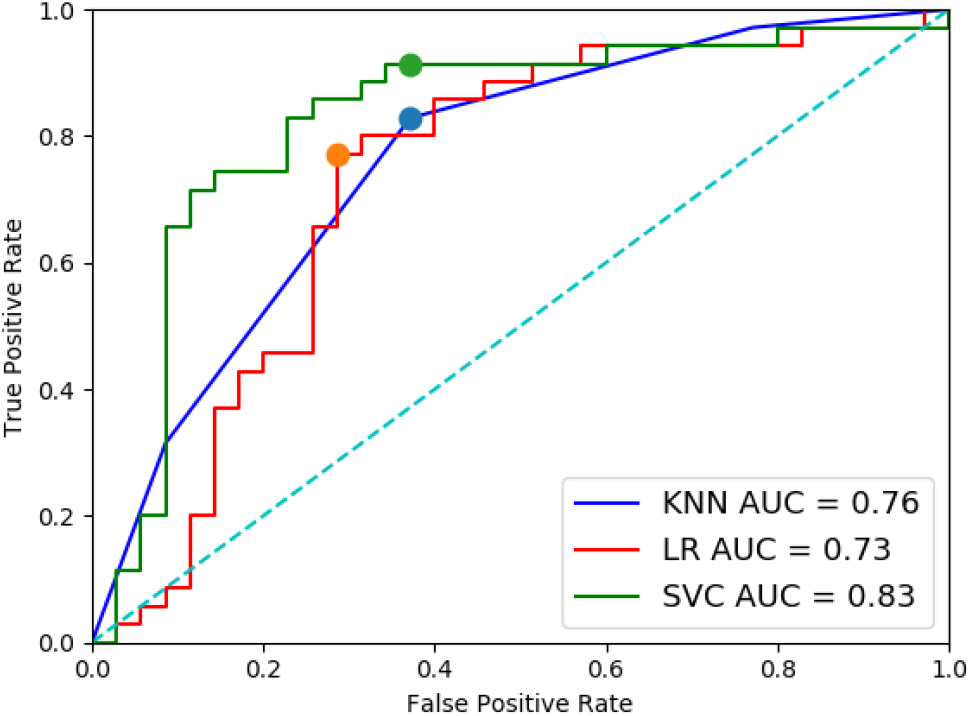
Receiver operating characteristic curves for the three classifiers evaluated in this work for cyanosis detection. The filled circles represent the operating points resulting in the other performance statistics. (Small differences exist due to the leave-one-out cross-validation approach.)

To assess the relative importance of the features used for classification, we visualized the weights assigned to different features by the logistic regression classifier, since this is easier to interpret than the parameters of the SVR or KNN. Figure 6 shows the weights for the eighteen features used (six bins each for each of the three R, G and B channels). The first six features correspond to the red channel (R1 - R6). The next six features correspond to the green channel (G1 - G6) and the last six features correspond to the blue channel (B1 - B6).

**Fig. 6.**
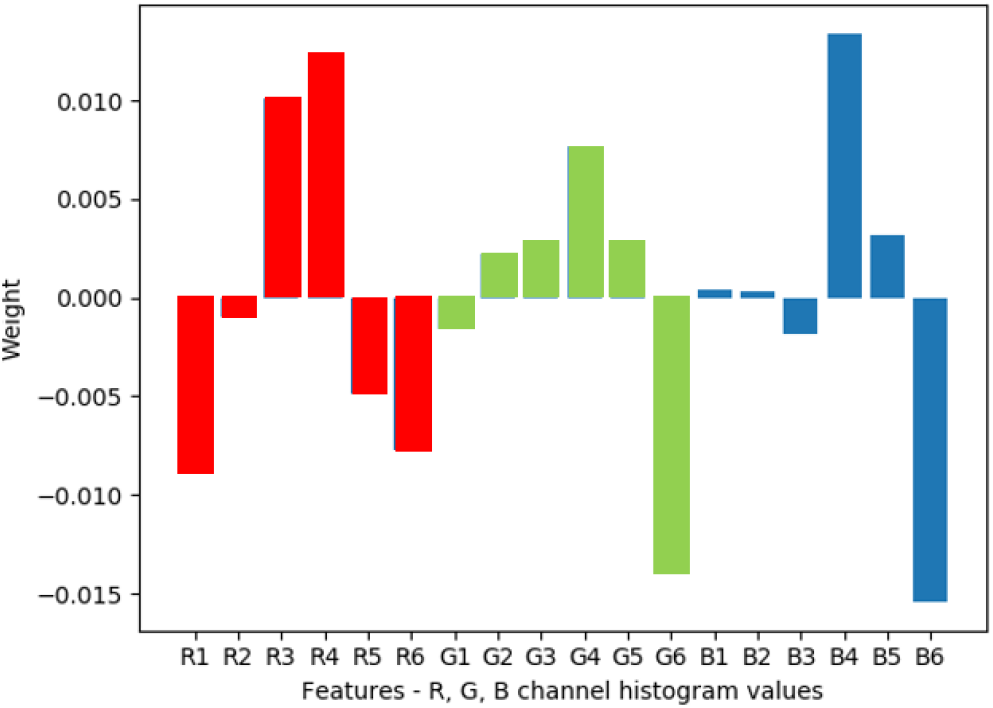
Weighting of features from logistic regression. The features are the histogram values the R, G and B channels. Positive coefficients refer to cyanotic condition and negative refer to non-cyanotic.

### R. Heart rate estimation

Figure 7 provides a Bland-Altman plot for the heart rate estimates. The mean of the absolute difference between the ground truth and estimate values is 16.31 bpm, and the standard deviation of this absolute difference is 14.42 bpm. It can be observed that the estimate struggles to provide an accurate hear rate above 70 bpm. This could be due to small movements within the ROI - the subject had to perform cardiovascular exercises to raise the heart rate. The breathing rate also increased, which caused movement in the face while inhaling and exhaling. Another issue that contributed to the errors was sudden changes in lighting.

**Fig. 7.**
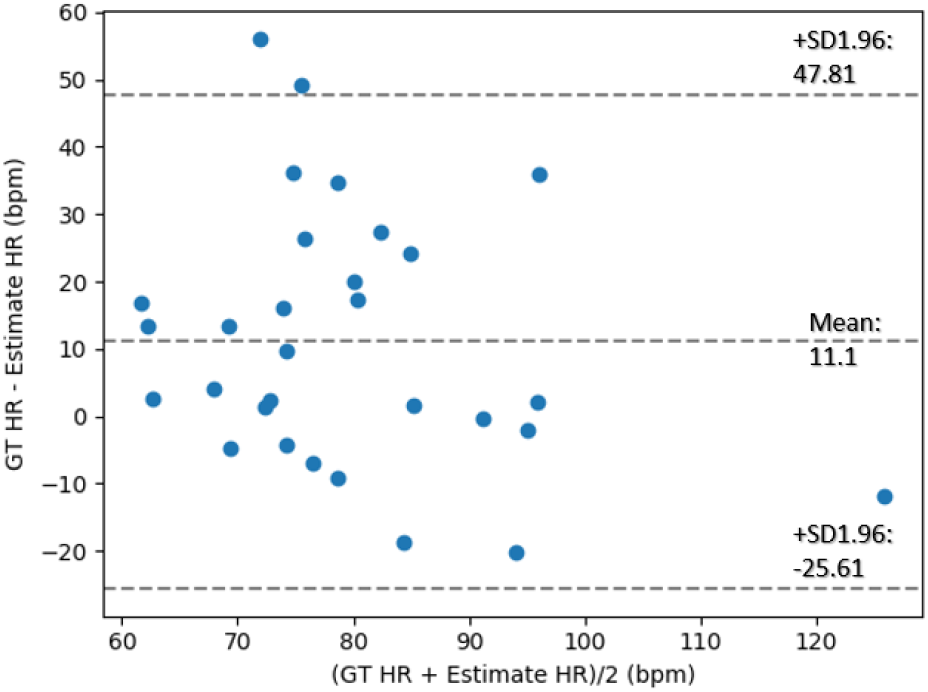
Bland-Altman plot of estimated heart rates from visible camera from a single subject

### S. Respiratory effort estimation

Figure 8 shows the time series signal obtained by the process described above (taking the mean value of all pixels of the difference between consecutive frames and passing them through a bandpass filter). On the left hand side is the signal obtained for rapid breathing (tachnypnea) at a respiratory rate of 20 breaths per minute. On the right side of the image is the signal obtained for hyperventilation at 60 breaths per minute.

**Fig. 8.**
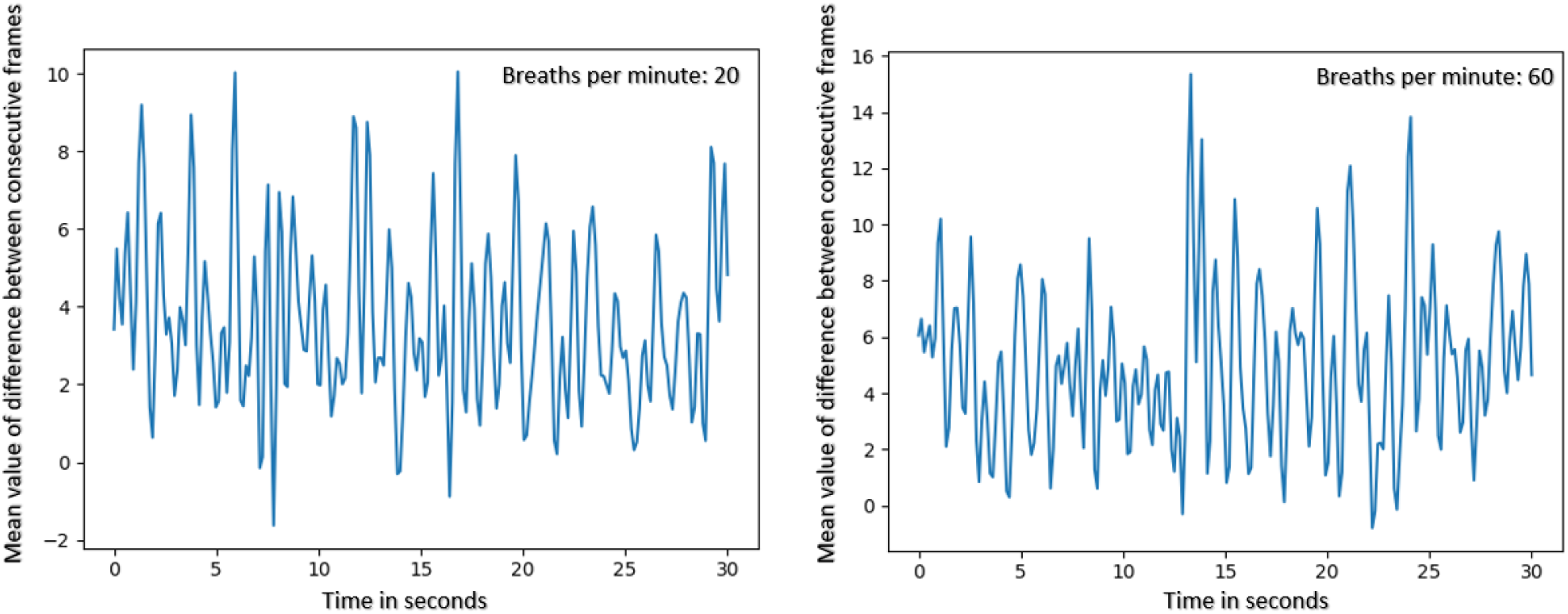
Plot of respiratory effort estimate over time at tow different rates

Figure 9 shows the distribution of energy in the 0.05Hz to 2Hz frequency range of the signal along side the corresponding respiratory rate measured. It can be seen that the energy values approximately track the respiratory rate. The Pearson correlation coefficient between the respiration rate and energy is 0.63.

**Fig. 9.**
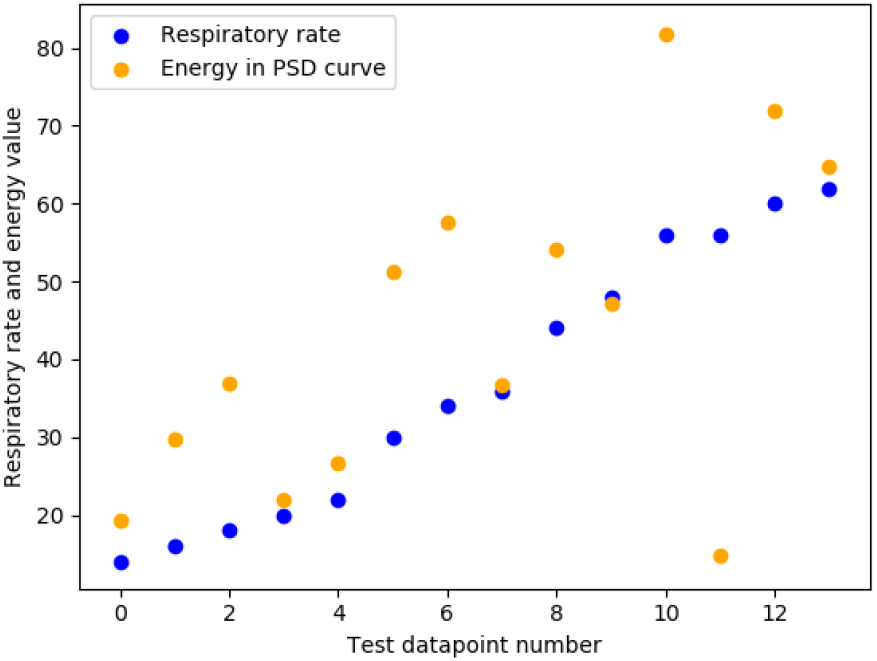
Plot of energy in signal against respiration rate.

### T. Clinical feedback

Figure 10 illustrates the system being used (hand-held version, not on tripod) in a field test at the emergency department. A series of informal tests for varying lighting conditions were made, and informal discussions were conducted with the clinical staff. (Formal testing was not possible because no ethical review approval had been made at the time.)

**Fig. 10.**
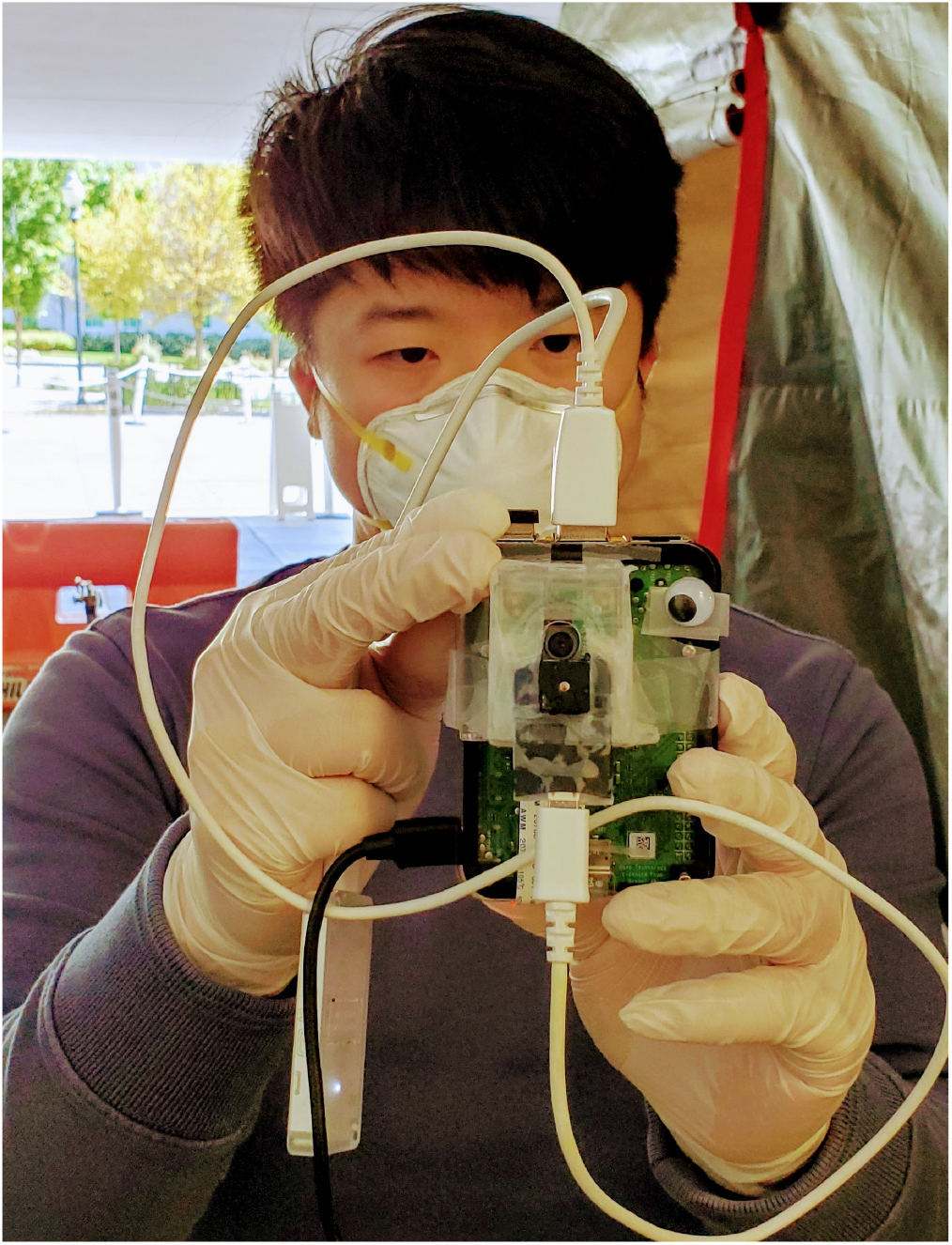
Photo of one of the authors testing out the system at the emergency department during a preliminary field trial.

## Discussion

### U. Forehead and lip segmentation

Detecting and segmenting out the forehead and lips is the first step in our pipeline. The accuracy of this stage can determine the accuracy of the remaining stages. This step is dependent on the performance of PoseNet, which sometimes provides false positive detection of individual’s faces. In this implementation, it is limited to detecting ten people in any given frame.

### V. Febrile state detection

Previous meta-reviews suggest that peripheral temperature may not be sufficient to determine fever (52, 53). This suggests that our proposed system, along with all traditional methods that measure peripheral temperature, like an ear thermometer, are not suitable to be used to perform an accurate diagnosis of febrile state. However, the proposed system is useful to perform mass early screening of the febrile state as a triage tool. The body temperature varies throughout the day in accordance with the circadian rhythm ((14), chapter 218). Taking this into account and having a dynamic threshold can reduce the number of false positive and false negative fever detection. Besides, body temperature could vary based on the ambient temperature. Having a reference temperature can help solve this issue. Additionally, the thermal camera in selection does not innately have the level of accuracy required for this task. Thus, a calibration for the targeted temperature range needs to be performed.

However, the temperature calibration of the thermal camera is not a trivial task and can be inaccurate depending on various aspects, like environmental temperature and surface condition. Also, a previous study suggests that improper use and interpretation of the infrared camera can lead to inaccurate triage (54). Hence, it is important to understand that the proposed system is only reliable for the designated task under limited conditions. For example, the presence of common cosmetics or clothing such as a turban or hijab can affect the accuracy of the estimated temperature from thermal camera (55). With a higher budget, the use of a thermal camera with higher accuracy and, if possible, one which is pre-calibrated can lead to a more reliable system. But a higher cost will inevitably lower the accessibility of the system.

### W. Cyanosis estimation

For the detection of cyanosis, out of the three classification approaches evaluated, the SVC exhibited the highest accuracy (77.1%), and KNN has the highest AUC (0.93), although this is not significantly higher than the AUC of 0.91 for the SVR. The SVR also produces the fewest false negatives (missed cyanosis), which at triage, is probably the most important feature of this system. For the open-source implementation, we, therefore, chose the SVR, although we note this is somewhat arbitrary at this point, given the size of the data set we used.

To visualize the effect of the features on the overall classification, we plot the LR coefficients for each feature (see figure 6).

It can be observed that the red channel exhibits a higher importance for cyanosis detection. When creating a histogram of the pixel values with six equally spaced bins for each of the three color channels, it can be seen that every channel (R, G, and B) contributes to the classification.

We note that the lip cyanosis dataset we used is relatively small and contains relatively good quality images. In the wild, the quality of images is not guaranteed to be high. This may be due to variations in lighting, occlusions, movement, angle of presentation, distances much greater than 1m, among other issues. Consistency of ambient lighting is an important factor in ensuring the correct classification of cyanosis (14). Camera parameters such as field of view and shutter speed can also influence the absolute color detected by the camera. Applying a color correction by using a color reference in the frame can solve this issue (56). Cosmetics applied to and around the lip can also interfere with the classification. In practice, we observed that if the mouth is open or teeth are visible, the cyanosis classifications tend to be inaccurate.

Perhaps the most important issue to consider is that of skin color and the variation of presentation of cyanosis across the human race. It has been reported that detection of peripheral cyanosis in individuals with distinct levels of cutaneous pigmentation can have different levels of difficulty, but can be considerably mitigated by selecting the appropriate skin site to perform the observations (57). Research into racial bias in facial recognition algorithms (44) also has highlighted just how dangerous it can be to use these algorithms out-of-the-box, without tuning to a population or thought about the bias it can create.

### X. Heart rate estimation

From the results obtained from our experiments, it can be seen that the algorithm is capable of estimating heart rate at low heart rates, but it requires further tuning in order to increase accuracy. The deviation from ground truth increased when there was a movement in the ROI and when there were large and frequent illumination changes, such as from a bright screen. Note that in this experiment, the subject was very still, and illumination changes in the environment were relatively low. This indicates a need for an improved heart rate estimation approach.

The algorithm is still susceptible to noise from lighting variation and unwanted movement, which affects the HR estimates. It is also seen that the heart rate tracking is less accurate as the heart rate increases, which may be due to movement in the ROI due to rapid breathing after performing cardiovascular exercises to increase the heart rate.

### Y. Respiratory effort estimation

Shortness of breath changes the breathing pattern when compared to normal breathing. This is evident from figure 8. Figure 9 shows that the energy in the respiratory effort signal is positively correlated with the respiratory rate measured. These results indicate that respiratory effort has the potential to be used in the classification of breathing as normal or abnormal.

Further studies will have to be conducted on a larger number of subjects with varied demographics in order to create an algorithm capable of performing this classification accurately. In the future, we aim to address issues that can cause interference with readings, such as lighting variation, movement of clothes, unwanted movement of the body, etc. This will make the algorithm more robust to noise.

### Z. Clinical testing

During a rapid group session, conducted mostly over video connections, and partially in person, we determined that the system would best be deployed in a mass triage situation. For this reason we chose to add a car battery supply. We found varying temperatures required a temperature sensor to help with calibration. Finally, clinical feedback stressed the need for heart rate and oxygen saturation assessment. (Saturations below 92% and heart rate above 110 beats per minutes were identified as reasons for triage through to the emergency department.) While heart rate was implementable in a short period of time, oxygen saturation was left for future work. We also chose to add respiratory effort estimation as a key vital sign, since its implementation is similar to that of heart rate estimation. Never-the-less, the lack of test subjects, and rapidity of turn-around in development, limited the accuracy of these additional vital signs.

## Conclusion

In this work, we have proposed a system that can detect fever and cyanosis and estimate heart rate and respiratory effort using a combination of visible light and thermal cameras operating on an edge computation platform that is running state-of-the-art deep learning algorithms. The system does not require any direct interaction between the device and either patients or healthcare workers. The source code needed to replicate our proposed system can be found on Github (39). It is important to note that PoseNet is image size and rotationally invariant (at least for most behaviors), and although we optimize the analysis to work at a 1m distance from the camera, this invariance should create robustness to movement to and from the camera, as well as within the frame. Many improvements can be made to this system to increase the classification performance and stability, including larger population studies and end-to-end deep learning. However, the need for *something* is acute and will be increasingly so in low resource areas.

Through this work, put together as a rapid response within a few days under lockdown, we hope to provide a starting point for automatic triage in clinical settings. Improving on this work could lead to novel implementations that may help streamline triage in clinics and hospitals, potentially during the current pandemic, where non-contact and rapid screening has distinct advantages for infection reduction.

## Data Availability

All data and code to repeat this work are available from:
DOI:10.5281/zenodo.3740776
https://github.com/cliffordlab/AutoTriage_release/

## Acknowledgments

This work was adapted from research funded by gifts from the Cox Foundation, Center for Discovery and Emory University.

## Supplementary Note A: Bill of Materials

**Table 3.** Parts required to assemble hardware required with retail prices at time of publication. ‡ indicates optional

| Part # | Part Name | Manufacturer | Price (\$) |
| --- | --- | --- | --- |
| 1 | Raspberry Pi 4 with 1-4 GB of RAM | Raspberry Pi | 61.20 |
| 2 | 16 GB microSD card | Sandisk | 5.99 |
| 3 | Google Coral USB accelerator | Google | 74.99 |
| 4 | Visible light (Red, Green and Blue; RGB) RasPi Camera v2 | Raspberry Pi | 27.50 |
| 5 | Temperature/Humidity sensor | Adafruit | 9.99 |
| 6 | FLIR Lepton 3.5 Radiometry Long-Wave Infrared Camera | FLIR | 200.00 |
| 7 | Purethermal-2 FLIR Lepton Smart I/O Module | FLIR | 100.00 |
| 8 ‡ | 3.5 inch Resistive Touch-Screen TFT Display | Jun-Electron | 22.99 |
| 9 ‡ | Car battery (12V, 44 Ah) | Optima | 199.99 |
| 10 ‡ | Top-Post Battery Cable Terminal Positive/Negative Clamp | BaiFM | 5.00 |
| 11 ‡ | 12V to 5V DC USB Step-Down Power Converter | Konnected | 12.99 |

## Notes

### Competing Interest Statement

The authors have declared no competing interest.

### Funding Statement

Much of the research is based on previous systems built under funding from the Cox Foundation and the Center for Discovery.

## Bibliography

1. Jesse M. Pines, Joshua A. Hilton, Ellen J. Weber, Annechien J. Alkemade, Hasan Al Shabanah, Philip D. Anderson, Michael Bernhard, Alessio Bertini, André Gries, Santiago Ferrandiz, Vijaya Arun Kumar, Veli-Pekka Harjola, Barbara Hogan, Bo Madsen, Suzanne Mason, Gunnar Öhlén, Timothy Rainer, Niels Rathlev, Eric Revue, Drew Richardson, Mehdi Sattarian, and Michael J. Schull. International Perspectives on Emergency Department Crowding. Academic Emergency Medicine, 18(12):1358–1370, 2011. ISSN 1553-2712. doi: 10.1111/j.1553-2712.2011.01235.x. ZSCC: NoCitationData[s0] https://onlinelibrary.wiley.com/doi/full/10.1111/j.1553-2712.2011.01235.x.

2. Claire Morley, Maria Unwin, Gregory M Peterson, Jim Stankovich, and Leigh Kinsman. Emergency department crowding: a systematic review of causes, consequences and solutions. PloS one, 13(8), 2018.

3. John C Moskop, Joel M Geiderman, Kenneth D Marshall, Jolion McGreevy, Arthur R Derse, Kelly Bookman, Norine McGrath, and Kenneth V Iserson. Another look at the persistent moral problem of emergency department crowding. Annals of emergency medicine, 74(3): 357–364, 2019.

4. Lindsey Woodworth. Swamped: Emergency department crowding and patient mortality. Journal of Health Economics, 70:102279, 2020.

5. M. Christien van der Linden, Barbara E. A. M. Meester, and Naomi van der Linden. Emergency department crowding affects triage processes. International Emergency Nursing, 29:27–31, November 2016. ISSN 1755-599X. doi: 10.1016/j.ienj.2016.02.003. https://www.sciencedirect.com/science/article/abs/pii/S1755599X16300052.

6. Dominik Aronsky, Ian Jones, Bill Raines, Robin Hemphill, Scott R Mayberry, Melissa A Luther, and Ted Slusser. An Integrated Computerized Triage System in the Emergency Department. AMIA Annu Symp Proc, 2008:16–20, 2008. ISSN 1942-597X. ZSCC: 0000046 https://www.ncbi.nlm.nih.gov/pmc/articles/PMC2656061/.

7. Tia Gao and David White. A next generation electronic triage to aid mass casualty emergency medical response. In 2006 International Conference of the IEEE Engineering in Medicine and Biology Society, volume Supplement, pages 6501–6504, August 2006. doi: 10.1109/IEMBS.2006.260881. ISSN: 1557-170X.

8. Roman Goldenberg, Dov Eilot, Grigory Begelman, Eugene Walach, Eyal Ben-Ishai, and Nathan Peled. Computer-aided simple triage (cast) for coronary ct angiography (ccta). International journal of computer assisted radiology and surgery, 7(6):819–827, 2012.

9. Amir Mirhaghi. Most patients are triaged using the emergency severity index. J. Nucl. Cardiol., 24(2):738–738, April 2017. ISSN 1071-3581, 1532-6551. doi: 10.1007/s12350-016-0704-z. ZSCC:0000000.

10. Canadian Agency for Drugs and Technologies in Health. Non-contact thermometers for detecting fever: a review of clinical effectiveness, 2014.

11. Krystal Bullers. Merck manuals. https://web.archive.org/web/20191001153051/https://www.merckmanuals.com/professional/infectious-diseases/biology-of-infectious-disease.

12. Merck Manual. Fever. https://web.archive.org/web/20191001153051/. https://www.merckmanuals.com/professional/infectious-diseases/biology-of-infectious-disease/fever Accessed: 2019-10-01.

13. Coronavirus (COVID-19) media information. https://newsroom.clevelandclinic.org/2020/04/02/coronavirus-covid-19-media-information/. Accessed: 2020-4-4.

14. Vivian L Clark and James A Kruse. Clinical methods: the history, physical, and laboratory examinations. Jama, 264(21):2808–2809, 1990.

15. Robert H. Shmerling. How’s your heart rate and why it matters? 2017.

16. Jouko Karjalainen and Matti Viitasalo. Fever and cardiac rhythm. Archives of internal medicine, 146(6):1169–1171, 1986.

17. Centers for Disease Control and Prevention. Symptoms of coronavirus. 2020.

18. JSM Peiris, S. Lai, LLM Poon, Y Guan, LYC Yam, W Lim, J Nicholls, WKS Yee, WW Yan, MT Cheung, et al. Coronavirus as a possible cause of severe acute respiratory syndrome. The Lancet, 361(9366):1319–1325, 2003.

19. Eddie Y.K Ng, G.J.L Kawb, and W.M Chang. Analysis of IR thermal imager for mass blind fever screening. Microvascular Research, 68(2):104–109, September 2004. ISSN 00262862. doi: 10.1016/j.mvr.2004.05.003. ZSCC: 0000158.

20. E.F.J. Ring, A. Jung, J. Zuber, P. Rutkowski, B. Kalicki, and U. Bajwa. Detecting Fever in Polish Children by Infrared Thermography. In Proceedings of the 2008 International Conference on Quantitative InfraRed Thermography. QIRT Council, 2008. doi: 10.21611/qirt.2008.03_07_17. ZSCC: NoCitationData[s0].

21. Armote Somboonkaew, Panintorn Prempree, Sirajit Vuttivong, Jutaphet Wetcharungsri, Supanit Porntheeraphat, Sataporn Chanhorm, Prasit Pongsoon, Ratthasart Amarit, Yuttana Intaravanne, Kosom Chaitavon, and Sarun Sumriddetchkajorn. Mobile-platform for automatic fever screening system based on infrared forehead temperature. In 2017 Opto-Electronics and Communications Conference (OECC) and Photonics Global Conference (PGC), pages 1–4, Singapore, July 2017. IEEE. ISBN 978-1-5090-6293-5. doi: 10.1109/OECC.2017.8114910. ZSCC: NoCitationData[s0].

22. Jia-Wei Lin, Ming-Hung Lu, and Yuan-Hsiang Lin. A Thermal Camera Based Continuous Body Temperature Measurement System. In 2019 IEEE/CVF International Conference on Computer Vision Workshop (ICCVW), pages 1681–1687, Seoul, Korea (South), October 2019. IEEE. ISBN 978-1-72815-023-9. doi: 10.1109/ICCVW.2019.00208. ZSCC: 0000000.

23. Nur Fatihah Binti Azmi, Frank Delbressine, Loe Feijs, and Sidarto Bambang Oetomo. Color correction of baby images for cyanosis detection. In Annual Conference on Medical Image Understanding and Analysis, pages 354–370. Springer, 2018.

24. Hiromitsu Kobayashi. Effect of measurement duration on accuracy of pulse-counting. Ergonomics, 56(12):1940–1944, 2013.

25. G. Balakrishnan, F. Durand, and J. Guttag. Detecting pulse from head motions in video. In 2013 IEEE Conference on Computer Vision and Pattern Recognition, pages 3430–3437, 2013.

26. M. A. Haque, R. Irani, K. Nasrollahi, and T. B. Moeslund. Heartbeat rate measurement from facial video. IEEE Intelligent Systems, 31(3):40–48, 2016.

27. Wim Verkruysse, Lars O Svaasand, and J Stuart Nelson. Remote plethysmographic imaging using ambient light. Optics express, 16(26):21434–21445, 2008.

28. Ming-Zher Poh, Daniel J McDuff, and Rosalind W Picard. Non-contact, automated cardiac pulse measurements using video imaging and blind source separation. Optics express, 18 (10):10762–10774, 2010.

29. Mohamed Abul Hassan, Aamir Saeed Malik, David Fofi, Naufal Saad, Babak Karasfi, Yasir Salih Ali, and Fabrice Meriaudeau. Heart rate estimation using facial video: A review. Biomedical Signal Processing and Control, 38:346–360, 2017.

30. Indu Ayappa, Robert G Norman, Ana C Krieger, Alison Rosen, Rebecca L O’Malley, and David M Rapoport. Non-invasive detection of respiratory effort-related arousals (reras) by a nasal cannula/pressure transducer system. Sleep, 23(6):763–771, 2000.

31. Leonardo Sarlabous, Abel Torres, Jose A Fiz, and Raimon Jane. Evidence towards improved estimation of respiratory muscle effort from diaphragm mechanomyographic signals with cardiac vibration interference using sample entropy with fixed tolerance values. PloS one, 9(2), 2014.

32. Andrew Bates, Martin J Ling, Janek Mann, and Damal K Arvind. Respiratory rate and flow waveform estimation from tri-axial accelerometer data. In 2010 International Conference on Body Sensor Networks, pages 144–150. IEEE, 2010.

33. Ciara O’Brien and Conor Heneghan. A comparison of algorithms for estimation of a respiratory signal from the surface electrocardiogram. Computers in biology and medicine, 37 (3):305–314, 2007.

34. Walter Karlen, Ainara Garde, Dorothy Myers, Cornie Scheffer, J Mark Ansermino, and Guy A Dumont. Estimation of respiratory rate from photoplethysmographic imaging videos compared to pulse oximetry. IEEE journal of biomedical and health informatics, 19(4):1331–1338, 2015.

35. Mark van Gastel, Sander Stuijk, and Gerard de Haan. Robust respiration detection from remote photoplethysmography. Biomedical optics express, 7(12):4941–4957, 2016.

36. Ehsaneh Shahhaidar, Ehsan Yavari, Jared Young, Olga Boric-Lubecke, and Cris Stickley. Respiratory effort energy estimation using doppler radar. In 2012 Annual International Conference of the IEEE Engineering in Medicine and Biology Society, pages 719–722. IEEE, 2012.

37. AP Prathosh, Pragathi Praveena, Lalit K Mestha, and Sanjay Bharadwaj. Estimation of respiratory pattern from video using selective ensemble aggregation. IEEE Transactions on Signal Processing, 65(11):2902–2916, 2017.

38. Christoph Hoog Antink, Simon Lyra, Michael Paul, Xinchi Yu, and Steffen Leonhardt. A broader look: Camera-based vital sign estimation across the spectrum. Yearbook of medical informatics, 28(01):102–114, 2019.

39. Chaitra Hegde and Zifan Jiang, Pradyumna Byappanahalli Suresha, and Gari D. Clifford. AutoTriage - An Open Source Raspberry Pi-based Clinical Screening System, April 2020.

40. George Papandreou, Tyler Zhu, Liang-Chieh Chen, Spyros Gidaris, Jonathan Tompson, and Kevin Murphy. Personlab: Person pose estimation and instance segmentation with a bottom-up, part-based, geometric embedding model. In The European Conference on Computer Vision (ECCV), September 2018.

41. Isoiso TR. 13154: Medical electrical equipment—deployment, implementation and operational guidelines for identifying febrile humans using a screening thermograph. International Organization for Standardization, 2009.

42. FLIR LEPTON® Engineering Datasheet. FLIR Systems, 3 2018. Lepton Engineering Datasheet, Rev: 200.

43. Fever temperatures: Accuracy and comparison. https://www.cigna.com/individuals-families/health-wellness/hw/medical-topics/fever-temperatures-tw9223. Accessed: 2020-4-4.

44. Joy Buolamwini and Timnit Gebru. Gender shades: Intersectional accuracy disparities in commercial gender classification. In Sorelle A. Friedler and Christo Wilson, editors, Proceedings of the 1st Conference on Fairness, Accountability and Transparency, volume 81 of Proceedings of Machine Learning Research, pages 77–91, New York, NY, USA, 23–24 Feb 2018. PMLR.

45. F. Pedregosa, G. Varoquaux, A. Gramfort, V. Michel, B. Thirion, O. Grisel, M. Blondel, P. Prettenhofer, R. Weiss, V. Dubourg, J. Vanderplas, A. Passos, D. Cournapeau, M. Brucher, M. Perrot, and E. Duchesnay. Scikit-learn: Machine learning in Python. Journal of Machine Learning Research, 12:2825–2830, 2011.

46. Xiaobai Li, Jie Chen, Guoying Zhao, and Matti Pietikainen. Remote heart rate measurement from face videos under realistic situations. In The IEEE Conference on Computer Vision and Pattern Recognition (CVPR), June 2014.

47. P. Viola and M. Jones. Rapid object detection using a boosted cascade of simple features. In Proceedings of the 2001 IEEE Computer Society Conference on Computer Vision and Pattern Recognition. CVPR 2001, volume 1, pages I–I, 2001.

48. Akshay Asthana, Stefanos Zafeiriou, Shiyang Cheng, and Maja Pantic. Robust discriminative response map fitting with constrained local models. In The IEEE Conference on Computer Vision and Pattern Recognition (CVPR), June 2013.

49. Haykin Simon. Adaptive filter theory. forth edition.–nj: by prentice-hall, 2002.

50. C. Li, C. Xu, C. Gui, and M. D. Fox. Distance regularized level set evolution and its application to image segmentation. IEEE Transactions on Image Processing, 19(12):3243–3254, 2010.

51. P. Welch. The use of fast fourier transform for the estimation of power spectra: A method based on time averaging over short, modified periodograms. IEEE Transactions on Audio and Electroacoustics, 15(2):70–73, 1967.

52. Bruce K Muma, David J Treloar, Karen Wurmlinger, Edward Peterson, and April Vitae. Comparison of rectal, axillary, and tympanic membrane temperatures in infants and young children. Annals of Emergency Medicine, 20(1):41–44, January 1991. ISSN 0196-0644. doi: 10.1016/S0196-0644(05)81116-3. ZSCC: 0000121 https://www.sciencedirect.com/science/article/abs/pii/S0196064405811163.

53. Binoy Mistry, Sarah Stewart De Ramirez, Gabor Kelen, Paulo S.K. Schmitz, Kamna S. Balhara, Scott Levin, Diego Martinez, Kevin Psoter, Xavier Anton, and Jeremiah S. Hinson. Accuracy and Reliability of Emergency Department Triage Using the Emergency Severity Index: An International Multicenter Assessment. Annals of Emergency Medicine, 71(5): 581–587.e3, May 2018. ISSN 01960644. doi: 10.1016/j.annemergmed.2017.09.036.

54. James B Mercer and E Francis J Ring. Fever screening and infrared thermal imaging: concerns and guidelines. Thermology International, 19(3):67–69, 2009.

55. Kaikai Zheng, Ruoyu Dong, Huan Wang, and Steve Granick. Infrared assessment of human facial temperature in the presence and absence of common cosmetics. preprint, Epidemiology, March 2020. ZSCC: 0000000.

56. Nur Fatihah Binti Azmi, Frank Delbressine, Loe Feijs, and Sidarto Bambang Oetomo. Color correction of baby images for cyanosis detection. In Mark Nixon, Sasan Mahmoodi, and Reyer Zwiggelaar, editors, Medical Image Understanding and Analysis, pages 354–370, Cham, 2018. Springer International Publishing. ISBN 978-3-319-95921-4.

57. Gladimir VG Baranoski, Spencer R Van Leeuwen, and Tenn F Chen. On the detection of peripheral cyanosis in individuals with distinct levels of cutaneous pigmentation. In 2017 39th Annual International Conference of the IEEE Engineering in Medicine and Biology Society (EMBC), pages 4260–4264. IEEE, 2017.

